# Fast and easy disinfection of coronavirus-contaminated face masks using ozone gas produced by a dielectric barrier discharge plasma generator

**DOI:** 10.1101/2020.04.26.20080317

**Authors:** Jinyeop Lee, Cheolwoo Bong, Pan K. Bae, Abdurhaman T. Abafogi, Seung H. Baek, Yong-Beom Shin, Moon S. Bak, Sungsu Park

## Abstract

Face masks are one of the currently available options for preventing the transmission of the severe acute respiratory syndrome coronavirus 2 (SARS-CoV-2), which has caused the 2019 pandemic. However, with the increasing demand for protection, face masks are becoming limited in stock, and the concerned individuals and healthcare workers from many countries are now facing the issue of the reuse of potentially contaminated masks. Although various technologies already exist for the sterilization of medical equipment, most of them are not applicable for eliminating virus from face masks. Thus, there is an urgent need to develop a fast and easy method of disinfecting contaminated face masks. In this study, using a human coronavirus (HCoV-229E) as a surrogate for SARS-CoV-2 contamination on face masks, we show that the virus loses its infectivity to a human cell line (MRC-5) when exposed for a short period of time (1 min) to ozone gas produced by a dielectric barrier discharge plasma generator. Scanning electron microscopy and particulate filtration efficiency (PFE) tests revealed that there was no structural or functional deterioration observed in the face masks even after they underwent excessive exposure to ozone (five 1-minute exposures). Interestingly, for face masks exposed to ozone gas for 5 min, the amplification of HCoV-229E RNA by reverse transcription polymerase chain reaction suggested a loss of infectivity under the effect of ozone, primarily owing to the damage caused to viral envelopes or envelope proteins. Ozone gas is a strong oxidizing agent with the ability to kill viruses on hard-to-reach surfaces, including the fabric structure of face masks. These results suggest that it may be possible to rapidly disinfect contaminated face masks using a plasma generator in a well-ventilated place.

## Introduction

Face masks are serving as one of the options for preventing severe acute respiratory syndrome coronavirus 2 (SARS-CoV-2), which has caused the 2019 international pandemic of the coronavirus disease.^1^ However, with increasing demands for protection, face masks are becoming limited in stock; concerned individuals and healthcare workers from many countries are now facing the issue of the reuse of potentially contaminated masks in many countries. Thus, there is an urgent need to develop a fast and easy method of disinfecting contaminated face masks. There already exist technologies for the sterilization of medical equipment, including personal protective equipment (PPE); these technologies include autoclave treatment, ethylene oxide gassing, ionized hydrogen peroxide fogging and hydrogen peroxide vaporization.^2^ However, most of them are not practical for disinfecting face masks with SARS-CoV-2.^2^ It was reported that ozone gas produced by plasma generators can inactivate various types of viruses on different surfaces, including porous ones.^3–6^ Ozone is a powerful oxidizing agent, but it does not linger. Its production, involving the use of electricity and a normal atmosphere, is easy and inexpensive. However, it has not been determined whether ozone gas can disinfect face masks contaminated with SARS-CoV-2 without compromising the filtration efficiency of the masks.

Here we show that a human coronavirus (HCoV-229E)^7,8^ as a surrogate for SARS-CoV-2 on face masks lost its infectivity to a human cell line (MRC-5) when exposed to gaseous ozone produced by a dielectric barrier discharge (DBD) plasma generator^9^ for a short time (1 min) (Figure 1). Neither structural nor functional deterioration of the face masks even with excessive exposures (5 times, 5 min per each time) to the ozone were observed by scanning electron microscopy (SEM) and a particulate filtration efficiency (PFE) test. Interestingly, RNA of HCoV-229E on the face masks by the ozone for 5 min was amplified by reverse transcription polymerase chain reaction (RT-PCR). This is the first demonstration of the potential of using ozone gas for disinfecting face masks contaminated with a coronavirus.

**Figure 1.**
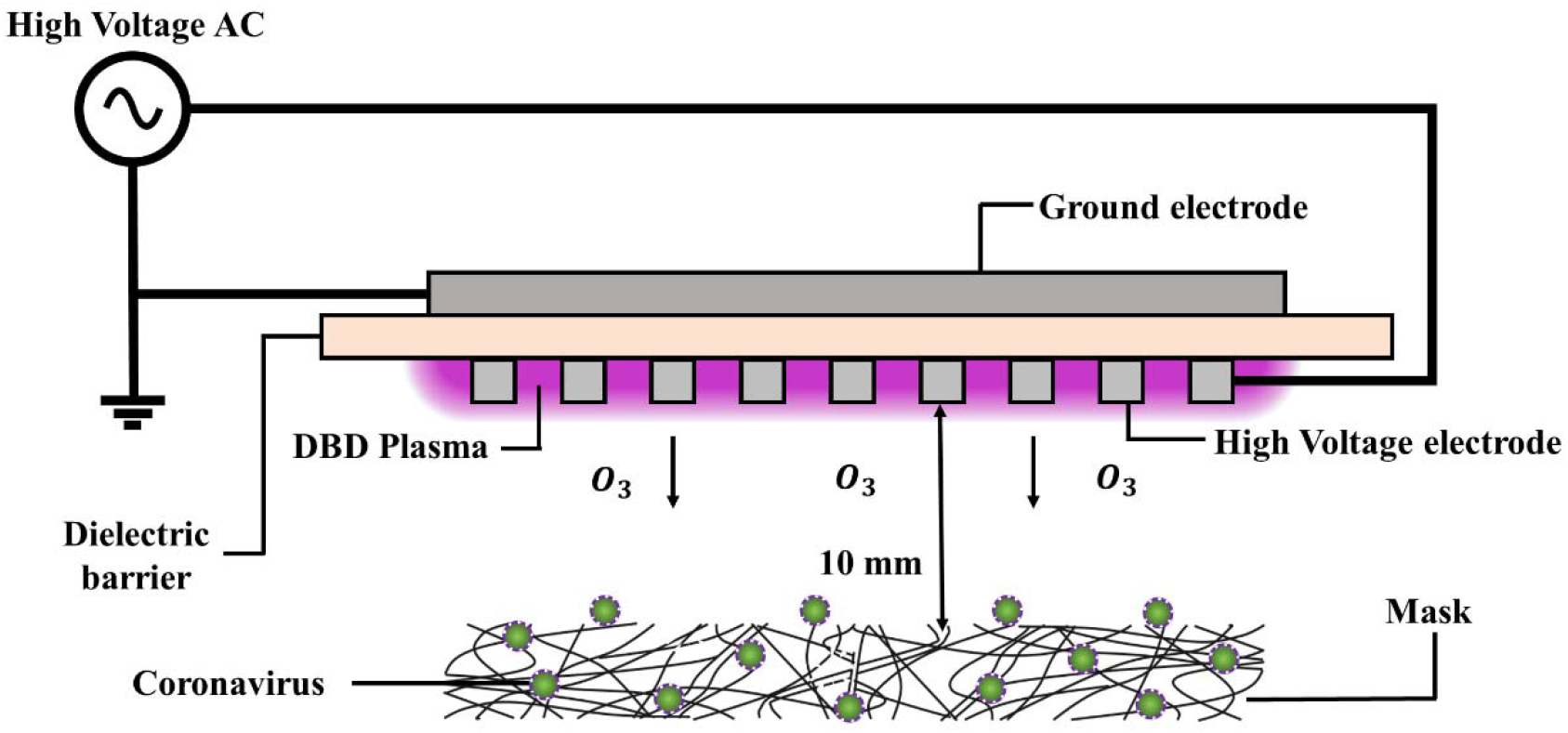
A schematic diagram describing the disinfection of a face mask contaminated by a coronavirus using ozone produced by a DBD plasma generator. It consists of a high-voltage, high-frequency power supply and two electrodes separated by a 1 mm-thick alumina dielectric barrier. Plasma was produced on the face of the device with the perforated electrode, along the rims of the holes.

## RESULTS AND DISCUSSION

### Inhibitory effect of ozone gas on virus and bacteria on face masks

Among the numerous types of plasma generators, the dielectric barrier discharge (DBD) plasma generator is considered the most energy-efficient and cost-effective plasma generator for ozone production; it forms ozone through the dissociation of molecular oxygen (O_2_) by collisions with excited electronic nitrogen populated by electron impacts and the ensuing combination between the atomic oxygen and O_2_.^9^

When face masks, experimentally contaminated with a human coronavirus (HCoV-229E)^7,8^ as a surrogate, were exposed to ozone gas (about 120 ppm) produced by the plasma generator for either 1 or 5 min, no viable HCoV-22E was recovered from the face masks (Table 1). Corresponding untreated face masks showed the recovery of about 3 log units of tissue culture infective dose 50% (log TCID_50_) per mL following 15 min of air drying. To the best of our knowledge, this is the first demonstration of the potential of using ozone gas for disinfecting face masks contaminated with a coronavirus. Similar results were obtained for face masks experimentally contaminated with either influenza A virus (H1N1)^10^ (Table S1) or Gram-positive bacteria *Staphylococcus aureus* (Table S2 and Figure S1) when exposed to ozone gas. These results suggest that virus and bacteria on face masks can be inactivated by ozone gas at a concentration of about 120 ppm within a short time (1-5 min).

**Table 1.** HCoV-229E titer recovered from contaminated face masks with and without exposure to ozone gas.

| Treatment of face masks<sup>a</sup> contaminated with HCoV-229E | Recovered virus<sup>b</sup><br>(log TCID<sub>50</sub> ± S.D.) |
| --- | --- |
| None | 3.0 ± 0.2 (n = 4) |
| Ozone gas (120 ppm<sup>c</sup>, 1 min) | 0 (n = 3) |
| Ozone gas (120 ppm, 5 min) | 0 (n = 3) |

<sup>a</sup> Samples (30 mm × 35 mm in size) were cut from face masks (Kleenguard®; product number Y2-44015, Kimberly-Clark Worldwide, Inc., Irving, TX, USA) with 3-layers filtering. The front side was sprayed with about 250 µL of HCoV-229E culture (about 4.5 log TCID<sub>50</sub> per mL) and dried at room temperature for 15 min in a biosafety cabinet before exposed to ozone gas.
<sup>b</sup> Virus particles on the samples were collected by washing the sample surface with 5 mL of PBS (phosphate buffered saline, pH 7.4) and measured using MRC-5 cells (ATCC, Bethesda, MD, USA).<sup>11</sup>
<sup>c</sup> The ozone concentrations produced by the DBD plasma generator were measured via UV absorption spectroscopy.<sup>12</sup>
n: sample number.

### Partial degradation of viral RNA by ozone gas

To understand the mechanism underlying the viral inactivation of face masks by ozone gas, the experimentally contaminated face masks (Kleenguard^®^; product # Y2-44015, Kimberly-Clark Worldwide, Inc., Irving, TX, USA)—with and without exposure to ozone gas for 5 min—were washed, and the washing solutions were assayed with the quantitative reverse transcription polymerase chain reaction (qRT-PCR).^13^ Surprisingly, there was no significant difference (*p* > 0.05; student’s t-test) in the amount of amplifiable RNAs between the unexposed and exposed masks, indicating that the short exposure may not fully degrade the viral RNA (Table 2). Similarly, the RNA of either H1N1 (Table S3) or *S. aureus* (Table S4) on the face masks was not totally degraded by the exposure to ozone gas. These results suggest that the loss of infectivity could be due to the damage to the viral envelope or envelope proteins, resulting in failure of the virus to attach itself to host cells.^14^

**Table 2:** qRT-PCR of HCoV-229E in washing solutions obtained from contaminated face masks with and without exposure to ozone gas.

| Treatment of face masks contaminated with HCoV-229E | Ct value<sup>a</sup><br>(mean ± S.D.) |
| --- | --- |
| None | 22.7 ± 0.4 (n = 6) |
| Ozone gas (120 ppm, 5 min) | 23.1 ± 0.6 (n = 6) |

<sup>a</sup> cycle threshold (Ct) is defined as the number of cycles required for the fluorescent signal to exceed the background signal level (threshold). n: sample number.

### No structural damage on the filter layer of face masks

To test if the exposure of face masks (Kleenguard^®^) to either plasma or ozone gas causes any damage to their filter layer, uncontaminated face masks were exposed to ozone gas for 5 min (five 1-minute exposures). We did not see any noticeable damage on the front and back side of the face masks with eyes and under a light microscope, either (data not shown). Their inner filter layer composed of polypropylene meltblown non-woven fabric was further examined under a SEM. As shown in Figure 2, there was no detectable structural damage caused to the filter layer of the exposed face masks. The result showed that the repeated exposures (5 times) of face masks to ozone gas did cause structural damage to the face masks.

**Figure 2.**
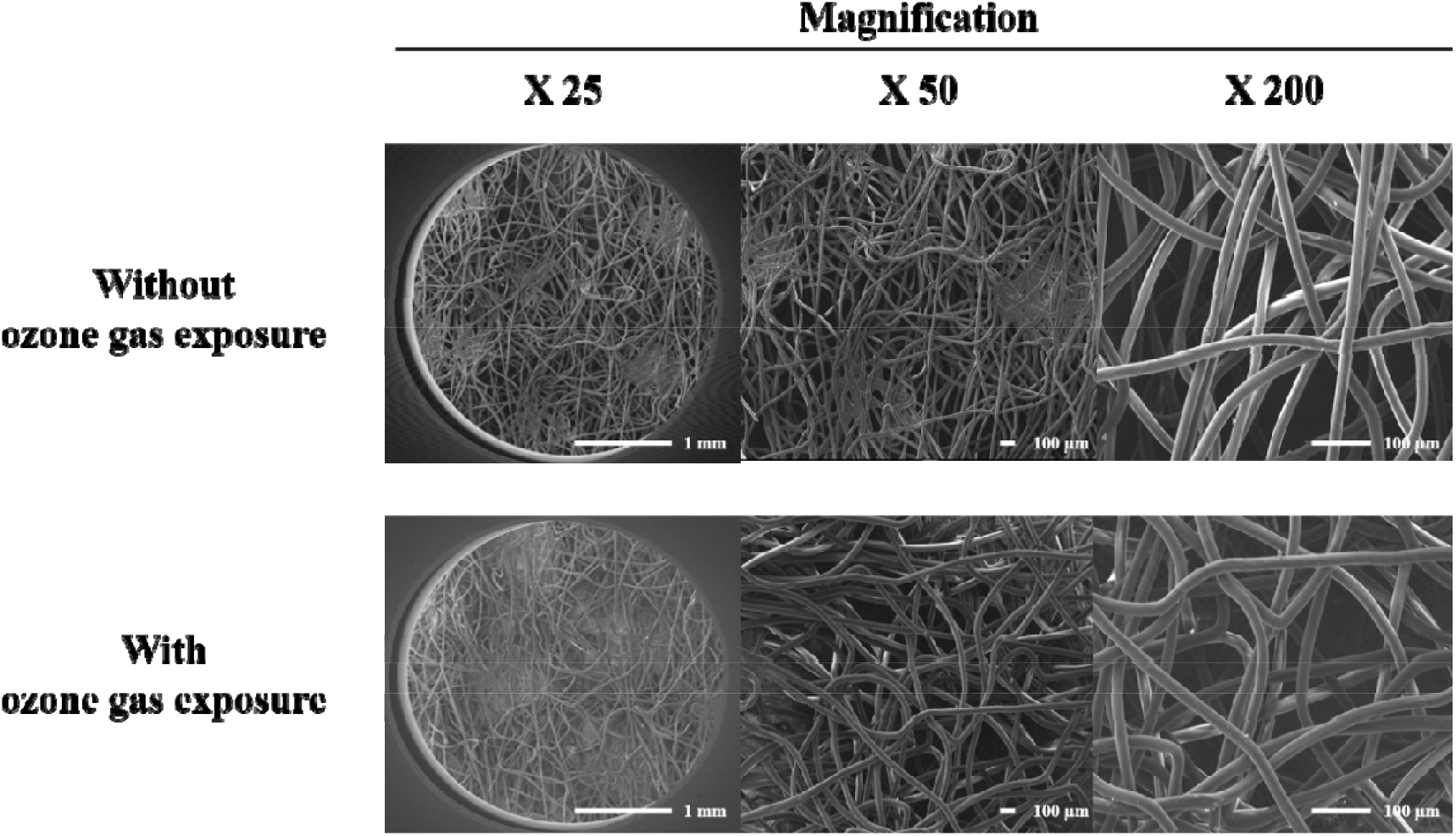
Scanning electron microscopy images of the filter layer of uncontaminated face masks (Kleenguard^®^) with and without exposures to ozone gas for 5 min (five 1-minute exposures). The images were taken by a field-emission scanning electron microscope (FE-SEM-EDS, JSM7500F, JEOL Ltd., Tokyo, Japan).

### No functional deterioration on face masks

The electrocharged filter is an essential component of dust masks such as N95 and KF94 masks and there was a concern that the charge on the filter could be lost with its exposure to ozone gas. The functioning of KF94 masks (registration number: F1-28712011; Kleannara Co., Seoul, Korea) which are certified to filter out 94% of particulate matter (about 0.4 μm diameter), after exposed to ozone gas for 5 min (five 1-minute exposures) was thus assessed using a standard test for measuring PFE with paraffin oil mists.^15^ There was no statistical difference in PFE between the exposed and unexposed face masks (Table 3). Taken together with the SEM images, it is suggested that the repeated exposures (5 times) of face masks to ozone gas do not cause structural or functional damage to the face masks.

**Table 3:** PFE of uncontaminated face masks (KF94) with and without exposure to ozone gas.

| Certified testing lab<sup>a</sup> | Ozone gas treatment on uncontaminated face masks | PFE (%)<sup>b</sup><br>(Mean ± S.D.) |
| --- | --- | --- |
| FITI Testing & Research Institute | None | 98.5 ± 1.3 (n=3) |
| | Ozone gas (120 ppm, 5 min) | 99.3 ± 1 (n=3) |
| Korea Mask Laboratory | None | 98.4 ± 0.5 (n=6) |
| | Ozone gas (120 ppm, 5 min) | 98.6 ± 0.5 (n=6) |

<sup>a</sup> The lab is certified and registered as a testing lab by the ministry of food and drug safety (MFDS) in Korea.
<sup>b</sup> Measured by a standard method using paraffin mist. n: sample number.

## CONCLUSIONS

In this study, using a human coronavirus (HCoV-229E) as a surrogate for SARS-CoV-2 contamination on face masks, the virus is shown to lose its infectivity to a human cell line (MRC-5) when exposed for a short period of time (1 min) to ozone gas produced by the DBD plasma generator. SEM and PFE tests revealed that there was no structural or functional deterioration observed in the face masks even after they underwent excessive exposure to ozone (five 1-minute exposures).

Ozone gas is a strong oxidizing agent with the ability to kill viruses on hard-to-reach surfaces, including the fabric structure of face masks. Inexpensive consumer-grade ozone generators are widely available. Our results suggest that it may be possible to rapidly disinfect face masks contaminated with SARS-CoV-2 using a plasma generator in a well-ventilated place.

## MATERIALS AND METHODS

### DBD plasma generator

The plasma generator consists of a high-voltage, high-frequency generator (Minipuls 2.2; GBS Elektronik GmbH, Großerkmannsdorf, Germany) and two electrodes separated by a 1 mm thick alumina dielectric barrier. Each electrode was made of a perforated stainless-steel plate and bare aluminum tape. The plasma was produced only at the perforated electrode, along the rims of the holes. As the dielectric barrier allows electrons and ions to accumulate on the surface, preventing the transition from a cold plasma to an arc, a sinusoidal voltage was applied to steadily produce the plasma (Figure S2). The plasma was turned on for 1 min after every 4 min to avoid any damage resulting from thermal heating.

### UV absorption spectroscopy

The ozone concentrations produced by the DBD plasma generator were measured via UV absorption spectroscopy.^16^ Light from a mercury lamp (BHK 90-0005-01, spectral line: 253.65 nm) was collimated using lenses and an optical fiber and sent through a gas medium, 4.3 cm above the electrode surface where the plasma is produced. The transmitted light intensity was then measured using a spectrometer (AvaSpec-2048L). As the wavelength-dependent absorption cross section of ozone is broad near 253.65 nm and known as 1.137 · 10^−17^ cm^2^molecule^−1^, the ozone concentration can be evaluated by the Beer-Lambert law described as follows:

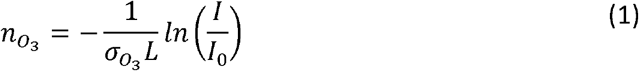

where *I* and *I*_0_ are the transmitted and incident light intensities; *n*_o3_ is the number density of ozone; 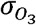 is the ozone absorption cross-section near 253.65 nm; *L* is the optical path length. The ozone concentrations under the test conditions were determined to be approximately 120 ppm.

### Coronavirus culture

HCoV-229E was cultured using human fetal lung fibroblast cell (MRC-5; ATCC, Bethesda, MD, USA) in a 96-well plate.^11^

### Exposure of face masks experimentally contaminated with HCoV-229E to ozone gas

Samples (n = 6) were cut (30 mm × 35 mm in size) from face masks and sprayed with about 250 μL of HCoV-229E culture (4.5 log TCID50 per mL) in a biosafety cabinet at a biosafety level-2 (BSL-2) laboratory while wearing face masks and gloves. The samples were then dried for 15 min at 25 °C and were individually exposed to ozone for both 1 and 5 min using the plasma generator (Figure 1) in a chemical hood.

### Determination of TCID_50_ value

Immediately after the exposure, the virus particles on the samples were collected by washing the samples with 5 mL of PBS (phosphate buffered saline, pH 7.4). TCID_50_ was determined by adding serial 10-fold dilutions of HCoV-229E collected from each mask into a human fetal lung fibroblast cell (MRC-5, ATCC, Bethesda, MD, USA) monolayer in a 96-well plate.^11^ The plates were observed for cytopathic effects for 4 days. The viral titer was calculated via the Reed and Munch endpoint method.^11^ Viral titer collected from the face masks was measured using MRC-5 cells (ATCC, Bethesda, MD, USA).^11^

### RT-PCR

Immediately after the exposure, the virus particles on the samples were collected by washing the samples with 5 mL of PBS. qRT-PCR was performed using StepOne™ Real-Time PCR system (Applied biosystems, CA, USA) and MG 2X One Step RT-PCR SYBR^®^ Green Master Mix reagents (Cancer Rop Co Ltd., Seoul, Korea). A segment of the *N* gene of HCoV229E was amplified using a forward primer (CGCAAGAATTCAGAACCAGAG) and a reverse primer (GGCAGTCAGGTTCTTCAACAA)^13^ with an amplicon size of 83 bp (Bioneer, Daejeon, Korea). The thermocycler conditions were as follows: reverse transcriptase at 50 °C for 30 min and an initial denaturation at 95 °C for 5 min, followed by 45 cycles of denaturation at 95 °C for 15 s, annealing at 52 °C for 30 s, and an extension at 72 °C for 30 s. To confirm that the target amplicon was properly formed, a melting curve analysis was conducted. The fluorescence intensity was measured within the range of 60–95 °C at a rate of 0.2 °C/s.

### SEM

A field-emission scanning electron microscope (FE-SEM-EDS, JSM7500F, JEOL Ltd., Tokyo, Japan) was used to take images of untreated and treated masks (n=4). To obtain the morphology of the electrostatic melt blown filter layer, which is the middle layer of the face mask, the melt blown filter was cut to obtain a 5 mm x 5 mm sample that was coated with iridium (Ir) for 15 minutes via the ion sputtering method. The FE-SEM-EDS was operated at 15 kV, and the working distance (WD) was 8 mm. All the sample images were acquired with magnification factors of 25, 50, and 200.

### Paraffin Oil Test

The standard test for measuring the filtration efficiency with paraffin oil mist was performed on masks at FITI Testing & Research Institute (Cheongju, Korea) and Korea Mask Laboratory (KML; Hanam city, Korea), the authorized testing organizations in Korea. In this test, a tester first produces paraffin oil mist with a particle size ranging from 0.05 to 1.7 μm, 0.4 μm on average.^15^ The flow containing the paraffin oil aerosol at a concentration of 20 ± 5 mg/m^3^ is then blown towards the mask at a flow rate of 95 L/min. The filtration efficiency, given by Eq. (2), is then evaluated by measuring the concentrations of paraffin oil mist upstream and downstream of the mask. The efficiency is a value averaged over 30 s and must be measured within 3 min after the test starts.

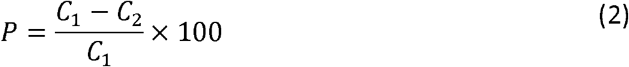

where *P* is the filtration efficiency; *C*_1_ and *C*_2_ are the concentrations of paraffin oil mist upstream and downstream of the mask, respectively.

## Data Availability

The data that support the findings of this study are available from the corresponding author, upon reasonable request.

## Supplementary Materials

Supplementary materials can be found at www.mdpi.com/xxx/s1.

## Acknowledgements

We thank Ms. J. So for giving an idea for the project.

## Funding

S.P. was supported by the BioNano Health-Guard Research Centre as a Global Frontier Project (H-guard 2018M3A6B2057299) through the National Research Foundation (NRF) of Ministry of Science and ICT (MSIT) in Korea. M.S.B. was supported by a grant through a future integration program of Kangbuk Samsung Hospital-Biomedical Institute for Convergence (BICS) at Sungkyunkwan University.

## Conflicts of Interest

The authors declare no conflict of interest.

## Notes

### Competing Interest Statement

The authors have declared no competing interest.

